# Hybrid humoral immune response of Pacific Islanders to BNT162b2 vaccination and Delta/Omicron infection: a cohort study

**DOI:** 10.1101/2024.04.09.24305559

**Authors:** Catherine Inizan, Adrien Courtot, Chloé Sturmach, Anne-Fleur Griffon, Antoine Biron, Timothée Bruel, Vincent Enouf, Thibaut Demaneuf, Sandie Munier, Olivier Schwartz, Ann-Claire Gourinat, Georges Médevielle, Marc Jouan, Sylvie van der Werf, Yoann Madec, Valérie Albert-Dunais, Myrielle Dupont-Rouzeyrol

## Abstract

**Background:** Pacific Islanders are underrepresented in vaccine efficacy trials. Few studies describe their immune response to COVID-19 vaccination. Yet, this characterization is crucial to re-enforce vaccination strategies adapted to Pacific Islanders singularities.

**Methods:** We evaluated the humoral immune response of 585 adults self-declaring as Melanesians, Europeans, Polynesians or belonging to other communities to Pfizer BNT162b2 vaccine. Anti-Spike and anti-Nucleoprotein IgG levels, capacity to neutralize SARS-CoV-2 variants and capacity to mediate Antibody-Dependent Cellular Cytotoxicity (ADCC) were assessed across communities at one and three months post-second dose or one and six months post-third dose.

**Results:** 61.3% of the sera tested contained anti-Nucleoprotein antibodies, evidencing mostly a hybrid immunity resulting from vaccination and SARS-CoV-2 infection. Anti-Spike IgG levels and capacity to mediate Omicron neutralization and ADCC were equivalent across the four ethnic communities at one-month post-immunization, during follow-up and at six months post-third dose, regardless of the infection status. Obese individuals (BMI>30 kg/m²) had significantly higher anti-Spike IgG levels at one-month post-immunization (+0.26 (0.04; 0.48) AU in LuLISA assay, *p* value = 0.017). Odds of Omicron neutralization at six months after the third dose decreased significantly in the 40-64 years and ≥65 years groups (OR (95% CI) 0.48 (0.24-0.90) and 0.29 (0.14-0.58) respectively, *p*-value = 0.003).

**Conclusions:** Our study evidenced Pacific Islander’s robust humoral immune response to Pfizer BNT162b2 vaccine, which is pivotal to re-enforce vaccination deployment in a population at risk for severe COVID-19 (clinicaltrials.gov: NCT05135585).

**summary:** Ethnicity has little impact on Pacific Islanders’ hybrid humoral immune response to BNT162b2 vaccination and SARS-CoV-2 infection. Anti-Spike IgG levels, capacity to neutralize Omicron variants and capacity to mediate Antibody-Dependent Cellular Cytotoxicity are equivalent across Pacific communities following BNT162b2 vaccination.

## Introduction

Covering 8.5 million km^2^ in a maritime area of 80 million km^2^, emerged Pacific territories excluding Australia are populated by ≈17 million inhabitants, among which 11.6 million belong to communities of Non-European Non-Asian descent ^1^. Pacific Islanders are at risk of severe respiratory infections, including influenza ^2^ and COVID-19 ^3^. As vaccination protects from severe COVID-19 in cohorts mostly composed of White people ^4^, it is pivotal to assess whether Pacific Islanders’ humoral response to anti-COVID-19 vaccination is equivalent to the one of White people.

New Caledonia (NC) is a French overseas territory of ≈270,000 inhabitants in the South Pacific region, where 41.2% self-declare as Melanesian, 24% as European, 8.3% as Polynesian and 26.5% as belonging to other communities ^5^.

A zero COVID policy protected NC from early SARS-CoV-2 spread. In September 2021, the first epidemic peak was caused by the Delta variant. Two larger epidemic peaks due to Omicron BA.1 and BA.4/.5 occurred in 2022 (Supplementary figure 1). Almost 80,000 infections and 314 deaths were detected ^6,7^. Vaccination roll-out, involving almost exclusively Pfizer BNT162b2 vaccine, was initiated in January 2021. As of May 2023, 70% of the entire population had received at least one dose, 68% had received at least two doses and 38% had received a booster dose ^8^.

Considering ethnic representation in COVID-19 vaccine trials is crucial ^9^. However, in Pfizer BNT162b2 efficacy trial, the cohort included only 0.2% Native Hawaiians/other Pacific Islanders ^10^, precluding to assess vaccine efficacy in this population. As a consequence, data on anti-SARS-CoV-2 immunity in vaccinated Pacific populations remain scarce. In a New Zealand cohort study, the 80 Pacific people included had lower anti-SARS-COV-2 antibody titers than Europeans and Maoris 28 days after two doses of BNT162b2 vaccine. However, when adjusting for the higher prevalence of diabetes in Pacific people, antibody titers and variant neutralization did not differ by ethnicity ^11^. Similarly, Australian First Nations people deployed a prototypical immune response to Pfizer BNT162b2 vaccine, with humoral and cellular responses at similar levels than Non-Indigenous people ^12^.

Anti-Spike antibody levels, neutralization capacity and antibodies functionality, including Antibody-Dependent Cellular Cytotoxicity (ADCC) ^13^, play an important role in COVID-19 pathogenesis. ADCC was indeed higher in hospitalized or severe patients who survived than in fatal cases ^13,14^, suggesting its protective role.

Adaptation of regional vaccination strategies to Pacific populations’ singularities requires to evaluate COVID-19 vaccines’ immunogenicity and hybrid immunity upon breakthrough infection in Pacific populations. The current study postulated that the hybrid humoral response to Pfizer BNT162b2 vaccine was equivalent across the four ethnic communities in NC. We aimed to characterize Pacific Islanders’ hybrid humoral response to BNT162b2 vaccine and SARS-CoV-2 infection, by evaluating anti-Spike IgG levels, the neutralization capacity and the ability to mediate ADCC of sera longitudinally collected in a cohort of 585 vaccinated adults from New Caledonia self-declaring as Melanesians, Polynesians, Europeans or belonging to other communities.

## Results

### Hybrid humoral response was highly prevalent in the cohort

This study enrolled 585 participants, among which 148 self-declared as Melanesians, 189 as Europeans, 82 as Polynesians and 166 as belonging to other communities. Sixty-eight participants were sampled at one month post-second dose (D2.1) and 56 participants were sampled at three months post-second dose (D2.3), of whom 50 participants were sampled at both timepoints (Figure 1). A total of 237 participants were sampled at 1 month post-third dose (D3.1) and 488 participants were sampled at 6 months post-third dose (D3.6), of whom 214 participants were sampled at both timepoints (Figure 1). Overall, 849 blood samples were collected and analyzed. All samples contained anti-Spike IgG.

**Figure 1.**
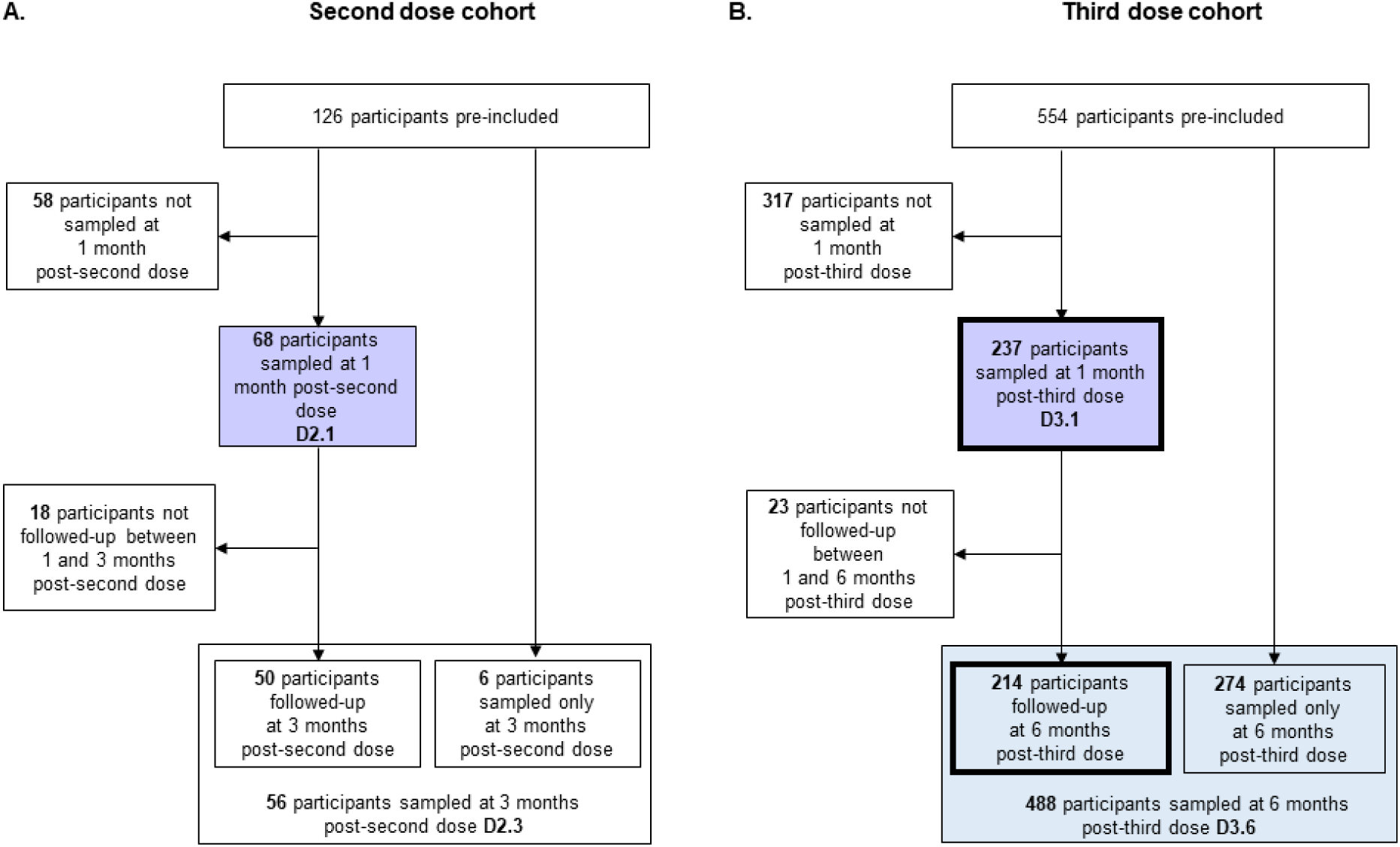
CONSORT (Consolidated Standards of Reporting Trials) diagrams of the second- and third-dose cohorts. **A.** The second dose cohort is composed of 126 pre-included participants, among which 68 were sampled one month after their second dose (purple square). **B.** The third-dose cohort is composed of 554 pre-included participants, among which 237 were sampled one month after their third dose (purple square), 214 were followed-up at one and six months after their third dose (bold frames) and 488 were sampled at 6 months after their third dose (blue square).

Eighty-five participants (14.5%) reported a positive COVID-19 test at one of the sampling timepoints. Participants exhibiting anti-Nucleocapsid antibodies, sign of a previous or current infection, at D2.1, D2.3, D3.1 and D3.6, were 47 (69.1%), 43 (76.8%), 150 (63.3%) and 280 (57.4%), respectively (Figure 2). In the 18 and 113 participants with anti-Nucleocapsid antibodies and followed up between D2.1/D2.3 or D3.1/D3.6, 9 re-infections and 9 new infections despite immunization (breakthrough infections) and 73 re-infections and 40 breakthrough infections were identified after the second and the third dose, respectively. Subsequent multivariate regression and logistic models were adjusted for infection status.

**Figure 2.**
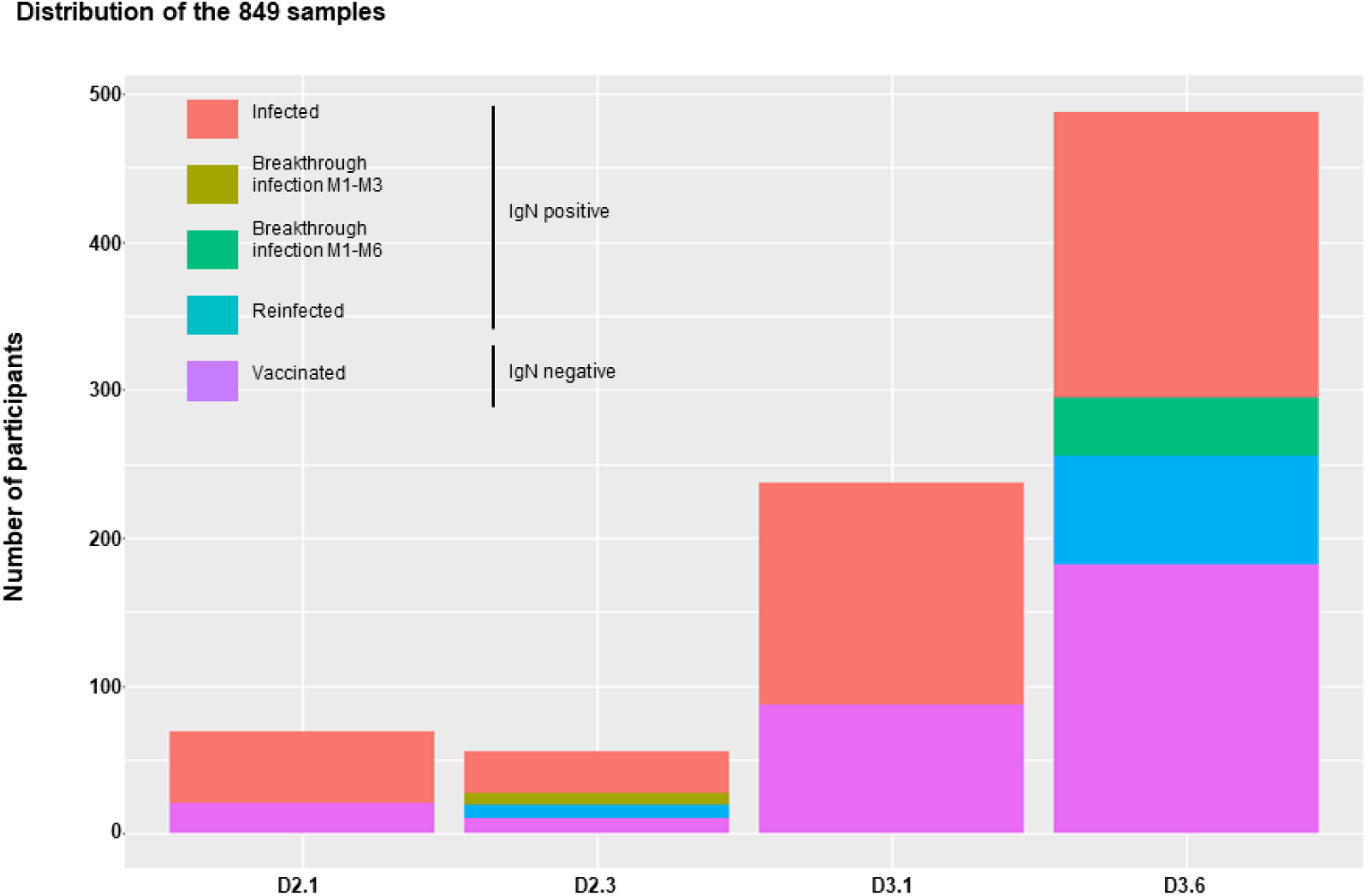
Description of the sample set. The 849 samples are divided into anti-N IgG (IgN) negative samples coming from vaccinated participants with no traces of previous infection (violet), and anti-N IgG positive samples coming from infected participants (red), breakthrough infections between one and three months post-second dose (brown), breakthrough infections between one and six months post-third dose (green) or reinfected participants (blue).

### Humoral response at one-month post-vaccination is equivalent across ethnic communities

Among the 305 participants sampled at one-month post-immunization (68 and 237 sampled at D2.1 and D3.1, respectively), ethnicity was distributed as follows: 55 Melanesians, 79 Europeans, 55 Polynesians and 116 participants belonging to other communities (Supplementary Table 1). The proportion of women was 54.6%. Age of participants ranged from 19 to 82 years, with European participants significantly older (*p*=0.002, Kruskal-Wallis). Previous infection was significantly less frequent in Europeans (*p*=0.005, Khi-2 test). BMI was significantly lower in Europeans (*p*<0.001, Kruskal-Wallis test).

Levels of anti-Spike IgG (anti-S IgG, *p*<0.001, Table 1 and Supplementary Table 2) and the ability to neutralize Omicron (*p*<0.001, Supplementary Table 2) were significantly higher one month after the third dose than after the second dose, irrespective of the infection status (Figure 3a). Previous SARS-CoV-2 infection was significantly associated with higher titers after the third dose (*p*<0.001, Figure 3A). After the second dose, Omicron neutralization was significantly more frequent in those with a previous infection (*p*=0.004, Table 2).

**Figure 3.**
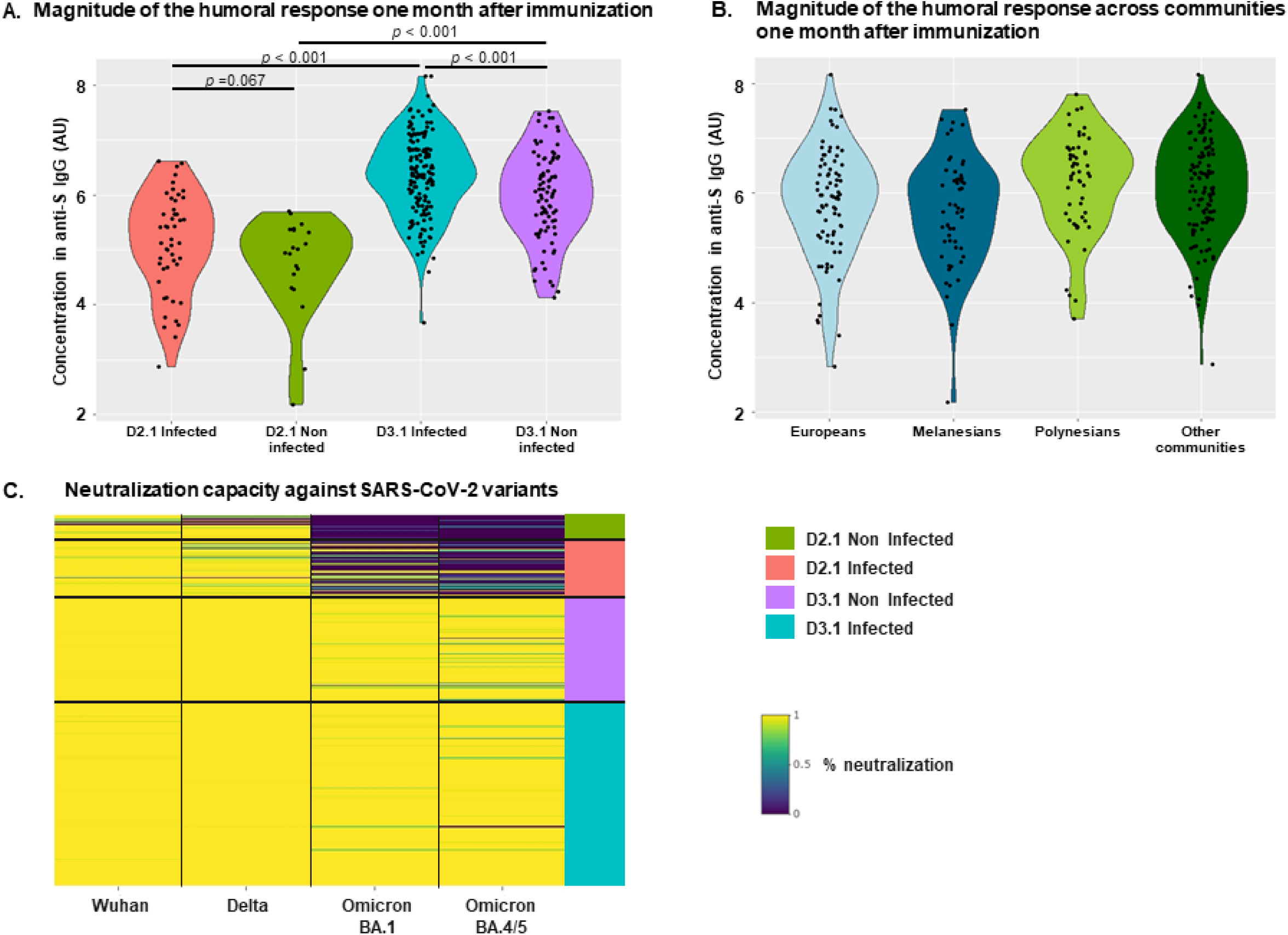
Measure of anti-S IgG levels and neutralization capacity. **A.** Concentrations in anti-S IgG in samples collected one month after the second dose from previously infected participants (D2.1 infected, red), one month after the second dose from previously non-infected participants (D2.1 non infected, green), one month after the third dose from previously infected participants (D3.1 infected, blue) and one month after the third dose from previously non-infected participants (D3.1 non infected, purple). **B.** Concentrations in anti-S IgG in samples collected from self-declared Melanesian (dark blue), European (light blue), Polynesian (light green) participants or from participants self-declaring as belonging to other communities (dark green) one month after immunization. **C.** Heatmap of the neutralization capacity against SARS-CoV-2 variants Wuhan, Delta, Omicron BA.1 and Omicron BA.4/5 one month after the second dose in previously non-infected participants (D2.1 non infected, green), one month after the second dose in previously infected participants (D2.1 infected, red), one month after the third dose in previously non-infected participants (D3.1 non infected, violet) and one month after the third dose in previously infected participants (D3.1 infected, blue). Gradient between dark blue and yellow denotes increasing neutralization capacity from 0% to 100% of 1/40 diluted sera. Each line represents one participant.

**Table 1.** Factors associated with anti-S IgG levels one month after immunization (linear regression)

|  | N=305 | Crude effect (95% CI) | p value | Adjusted effect (95% CI) | p value |
| --- | --- | --- | --- | --- | --- |
| **Timepoint** |  |  | **<0.001** |  | **<0.001** |
| **Post 2<sup>nd</sup> dose** | 68 | *Reference* |  | *Reference* |  |
| **Post 3<sup>rd</sup> dose** | 237 | **+1.24 (1.02; 1.46)** |  | **+1.27 (1.06; 1.49)** |  |
| **Infected** |  |  | **0.004** |  | **<0.001** |
| **No** | 108 | *Reference* |  | *Reference* |  |
| **Yes** | 197 | **+0.33 (0.11; 0.56)** |  | **+0.38 (0.19; 0.56)** |  |
| **Gender<sup>a</sup>** |  |  | **0.051** |  | **0.036** |
| **Male** | 138 | -0.18 (-0.36; 0.001) |  | **-0.19 (-0.37; -0.01)** |  |
| **Female** | 167 | *Reference* |  | *Reference* |  |
| **Age (years)<sup>a</sup>** |  |  | **0.046** |  |  |
| **18-39** | 130 | *Reference* |  |  |  |
| **40-64** | 133 | -0.14 (-0.34; 0.06) |  |  |  |
| **≥65** | 42 | **-0.35 (-0.63; -0.06)** |  |  |  |
| **Comorbidities<sup>a</sup>** |  |  | 0.51 |  |  |
| **No** | 173 | *Reference* |  |  |  |
| **Yes** | 132 | -0.06 (-0.25; 0.12) |  |  |  |
| **BMI<sup>a</sup>** |  |  | **0.017** |  | **0.008** |
| **Underweight** | 9 | -0.26 (-0.80; 0.29) |  | -0.48 (-1.05; 0.10) |  |
| **Normal** | 93 | *Reference* |  | *Reference* |  |
| **Overweight** | 90 | -0.04 (-0.27; 0.19) |  | +0.05 (-0.28; 0.18) |  |
| **Obese** | 113 | **+0.26 (0.04; 0.48)** |  | **+0.24 (0.03; 0.46)** |  |
| **Community<sup>a</sup>** |  |  | 0.17 |  |  |
| **European** | 79 | *Reference* |  |  |  |
| **Melanesian** | 55 | +0.02 (-0.26; 0.30) |  |  |  |
| **Polynesian** | 55 | +0.24 (-0.04; 0.52) |  |  |  |
| **Other** | 116 | +0.20 (-0.03; 0.44) |  |  |  |

<sup>a</sup>The univariate analysis is adjusted for timepoint and infection status.
CI: confidence interval; BMI: body mass index.
BMI classes: Underweight = BMI < 18.5 kg/m<sup>2</sup>, Normal weight = BMI ∈ [18.5, 25[ kg/m<sup>2</sup>, Overweight = BMI ∈ [25, 30[ kg/m<sup>2</sup>, Obese = BMI ≥ 30 kg/m<sup>2</sup>
Statistically significant associations (p < 0.05) with anti-S IgG levels are shown in bold.

**Table 2.** Factors associated with the ability to neutralize Omicron BA.1 and/or BA.4-5 one month after the second and third dose (logistic regression)

| *One month after the second dose* |  |  |  |  |
| --- | --- | --- | --- | --- |
|  | N=68 | Neutralisation, n (%) | Crude OR (95% CI) | p value |
| **Infected<sup>a</sup>** |  |  |  | **0.004** |
| No | 21 | 0 (0) | 1 |  |
| Yes | 47 | 13 (27.7) | **16.83 (2.04-2.19)** |  |
| **Level of anti-S IgG** |  |  |  | **<0.001** |
| <5.737 AU | 56 | 3 (5.4) | 1 |  |
| ≥ 5.737 AU | 12 | 10 (83.3) | **88.33 (15.79-805.92)** |  |
| **Gender** |  |  |  | 0.60 |
| Male | 31 | 5 (16.1) | 0.70 (0.19-2.36) |  |
| Female | 37 | 8 (21.6) | 1 |  |
| **Age (years)<sup>a</sup>** |  |  |  | 0.69 |
| 18-39 | 42 | 9 (21.4) | 1 |  |
| 40-64 | 20 | 4 (20.0) | 0.96 (0.25-3.30) |  |
| ≥65 | 6 | 0 (0) | 0.27 (0.00-2.67) |  |
| **Comorbidities** |  |  |  | 0.60 |
| No | 38 | 8 (21.1) | 1 |  |
| Yes | 30 | 5 (16.7) | 0.75 (0.20-2.54) |  |
| **BMI** |  |  |  | **0.14** |
| Normal weight | 20 | 2 (10) | 1 |  |
| Overweight | 23 | 3 (13.0) | 1.35 (0.20-11.14) |  |
| Obese | 25 | 8 (32) | 4.24 (0.90-30.81) |  |
| **Community** |  |  |  | 0.59 |
| European | 16 | 3 (18.8) | 1 |  |
| Melanesian | 22 | 3 (13.6) | 0.68 (0.11-4.20) |  |
| Polynesian | 9 | 1 (11.1) | 0.54 (0.02-5.11) |  |
| Other | 21 | 6 (28.6) | 1.73 (0.38-9.54) |  |
| *One month after the third dose* |  |  |  |  |
|  | N=237 | Neutralisation, n (%) | Crude OR (95% CI) | p value |
| **Infected** |  |  |  | 0.30 |
| No | 87 | 84 (96.6) | 1 |  |
| Yes | 150 | 148 (98.7) | 2.64 (0.43-20.36) |  |
| **Level of anti-S IgG** |  |  |  | **0.028** |
| <5.737 AU | 62 | 58 (93.5) | 1 |  |
| ≥ 5.737 AU | 175 | 174 (99.4) | **12.00 (1.73-237.30)** |  |
| **Gender** |  |  |  | 0.50 |
| Male | 130 | 128 (98.5) | 0.54 (0.07-3.33) |  |
| Female | 107 | 104 (97.2) | 1 |  |
| **Age (years)<sup>a</sup>** |  |  |  | 0.40 |
| 18-39 | 88 | 88 (100) | 1 |  |
| 40-64 | 113 | 109 (96.5) | 0.14 (0.00-1.32) |  |
| ≥65 | 36 | 35 (97.2) | 0.13 (0.00-2.57) |  |
| **Comorbidities** |  |  |  | 0.40 |
| No | 135 | 133 (98.5) | 1 |  |
| Yes | 102 | 99 (97.1) | 0.50 (0.06-3.05) |  |
| **BMI** |  |  |  | 0.32 |
| Underweight | 8 | 7 (87.5) | 0.19 (0.02-4.49) |  |
| Normal weight | 74 | 72 (97.3) | 1 |  |
| Overweight | 67 | 66 (98.5) | 1.83 (0.17-40.00) |  |
| Obese | 88 | 87 (98.9) | 2.42 (0.23-52.63) |  |
| **Community<sup>a</sup>** |  |  |  | 0.60 |
| **European** | 63 | 61 (96.8) | 1 |  |
| **Melanesian** | 33 | 33 (100) | 2.72 (0.21-379.95) |  |
| **Polynesian** | 46 | 44 (95.7) | 0.72 (0.11-4.85) |  |
| **Other** | 95 | 94 (98.9) | 2.56 (0.33-28.48) |  |

*CI: confidence interval; BMI: body mass index, OR: Odds Ratio.*
*<sup>a</sup>Logistic regression with Firth's correction.*
*BMI classes: Underweight = BMI < 18.5 kg/m<sup>2</sup>, Normal weight = BMI ∈ [18.5, 25[ kg/m<sup>2</sup>, Overweight*
*= BMI ∈ [25, 30[ kg/m<sup>2</sup>, Obese = BMI ≥ 30 kg/m<sup>2</sup>*
*Statistically significant associations (p < 0.05) with the ability to neutralize Omicron BA.1 and/or*
*BA.4-5 are shown in bold.*

Linear regression confirmed that number of doses and infection status were independently and significantly associated with higher anti-S IgG titers (adjusted effect +1.27, 95% CI (1.06; 1.49), *p*<0.001 and + 0.38, 95% CI (0.19; 0.56), *p*<0.001, respectively, Table 1). Obesity was associated with significantly higher anti-S IgG titers (adjusted effect +0.24, 95% CI (+0.03; +0.46), *p*=0.008). Male gender was significantly associated with lower anti-S IgG titers (adjusted effect -0.19, 95% CI (-0.37; - 0.01), *p*=0.036). Importantly, ethnic community was not associated with anti-S IgG titers (*p*= 0.34) (Figure 3B).

In a ROC curve analysis, a threshold of 5.737 AU maximized the proportion of subjects exhibiting anti-S IgG levels above the threshold who were able to neutralize Omicron (75% sensitivity) while minimizing the proportion of participants below the threshold able to neutralize Omicron (95% specificity) (Supplementary figure 2).

Pseudovirus neutralization assays showed that almost all individuals sampled one month after immunization were able to neutralize the vaccine strain Wuhan and the Delta variant (Figure 3C). The ability to neutralize Omicron BA.1 and/or BA.4/5 however significantly differed by number of doses (Figure 3C), with 19.1% and 97.9% participants neutralizing Omicron after the second and third dose, respectively (Supplementary Table 2). In analyses stratified by immunization dose, only anti-S IgG levels were significantly associated with neutralization (OR (95% CI): 88.33 (15.79-805.92), *p*<0.001 at D2.1, and 12.00 (1.73-237.30), *p*=0.028 at D3.1 Table 2). After two immunizations, only anti-S IgG levels remained significantly associated with Omicron neutralization (*p*<0.001). After three immunizations, in univariate analysis, no factor other than anti-S IgG levels (*p*=0.028) was associated with Omicron neutralization (all *p*-values>0.20). Importantly, ethnic community was not associated with different odds to neutralize Omicron *(p*=0.59 and *p*=0.60 at D2.1 and D3.1, respectively).

All participants but 7 were able to activate CD16 following exposure to SARS-CoV-2 Spike, reflecting a capacity to mediate ADCC. Multivariate linear regression showed that anti-S IgG levels ≥ 5.737 AU and male gender were independently associated with higher ADCC activation (+0.61 (0.29; 0.94), *p*<0.001 and +0.47 (0.14; 0.79), *p*=0.005, respectively, Table 3). Importantly, ethnic community was not associated with the capacity to mediate ADCC (*p*= 0.069 in multivariate linear regression).

**Table 3.**
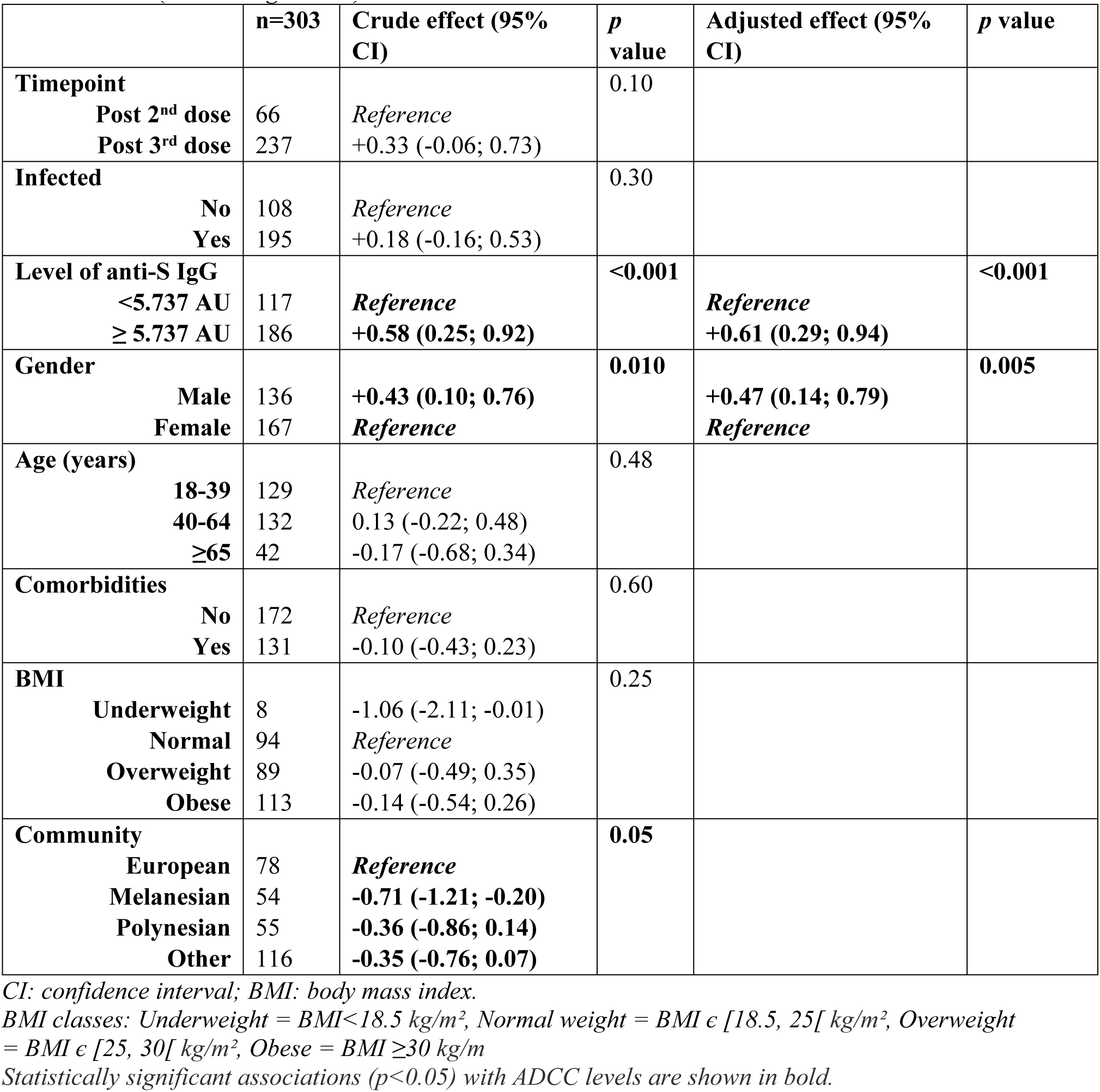
Factors associated with the level of ADCC (CD16 activation) one month after immunization (Linear regression)

|  | n=303 | Crude effect (95% CI) | p value | Adjusted effect (95% CI) | p value |
| --- | --- | --- | --- | --- | --- |
| **Timepoint** |  |  | 0.10 |  |  |
| **Post 2<sup>nd</sup> dose** | 66 | *Reference* |  |  |  |
| **Post 3<sup>rd</sup> dose** | 237 | +0.33 (-0.06; 0.73) |  |  |  |
| **Infected** |  |  | 0.30 |  |  |
| **No** | 108 | *Reference* |  |  |  |
| **Yes** | 195 | +0.18 (-0.16; 0.53) |  |  |  |
| **Level of anti-S IgG** |  |  | <0.001 |  | <0.001 |
| **<5.737 AU** | 117 | *Reference* |  | *Reference* |  |
| **≥ 5.737 AU** | 186 | **+0.58 (0.25; 0.92)** |  | **+0.61 (0.29; 0.94)** |  |
| **Gender** |  |  | 0.010 |  | 0.005 |
| **Male** | 136 | **+0.43 (0.10; 0.76)** |  | **+0.47 (0.14; 0.79)** |  |
| **Female** | 167 | *Reference* |  | *Reference* |  |
| **Age (years)** |  |  | 0.48 |  |  |
| **18-39** | 129 | *Reference* |  |  |  |
| **40-64** | 132 | 0.13 (-0.22; 0.48) |  |  |  |
| **≥65** | 42 | -0.17 (-0.68; 0.34) |  |  |  |
| **Comorbidities** |  |  | 0.60 |  |  |
| **No** | 172 | *Reference* |  |  |  |
| **Yes** | 131 | -0.10 (-0.43; 0.23) |  |  |  |
| **BMI** |  |  | 0.25 |  |  |
| **Underweight** | 8 | -1.06 (-2.11; -0.01) |  |  |  |
| **Normal** | 94 | *Reference* |  |  |  |
| **Overweight** | 89 | -0.07 (-0.49; 0.35) |  |  |  |
| **Obese** | 113 | -0.14 (-0.54; 0.26) |  |  |  |
| **Community** |  |  | 0.05 |  |  |
| **European** | 78 | *Reference* |  |  |  |
| **Melanesian** | 54 | **-0.71 (-1.21; -0.20)** |  |  |  |
| **Polynesian** | 55 | **-0.36 (-0.86; 0.14)** |  |  |  |
| **Other** | 116 | **-0.35 (-0.76; 0.07)** |  |  |  |

CI: confidence interval; BMI: body mass index.
BMI classes: Underweight = BMI < 18.5 kg/m<sup>2</sup>, Normal weight = BMI ∈ [18.5, 25[ kg/m<sup>2</sup>, Overweight = BMI ∈ [25, 30[ kg/m<sup>2</sup>, Obese = BMI ≥ 30 kg/m<sup>2</sup>
Statistically significant associations (p < 0.05) with ADCC levels are shown in bold.

### Humoral response to SARS-CoV-2 over 6 months post-third dose is similar across ethnic communities

Among the 214 participants who were evaluated at both D3.1 and D3.6, 29 self-declared as Melanesians, 57 as Europeans, 42 as Polynesians and 86 as belonging to other communities (Supplementary table 3). Europeans were significantly older (*p*=0.022, Kruskal-Wallis test), showed less frequently signs of previous infection (*p*=0.059, Khi² test) and had lower BMI (*p*<0.001, Kruskal-Wallis test) (Supplementary table 3). Between D3.1 and D3.6, anti-S IgG levels decreased in median (IQR) by 1.67 AU (0.63; 2.59) (Supplementary table 3). The multivariate linear regression model showed that in participants never infected and with anti-S IgG levels <5.737 at D3.1, the anti-S IgG levels decreased (in mean (95% CI)) by 1.98 AU (1.56; 2.39) between D3.1 and D3.6 (Supplementary table 4). In participants with anti-S IgG levels >5.737 at D3.1, the decrease was significantly steeper (-2.52 AU (-2.90; -2.14), *p*<0.001). In participants with breakthrough infection, anti-S IgG levels remained stable.

Two hundred and nine participants out of 214 showed the ability to neutralize Omicron at D3.1 (Supplementary table 3). Of them, 35 (16.7%) had lost this ability by D3.6 (Supplementary table 3). Multivariate logistic regression evidenced that previously infected individuals had significantly lower odds to lose Omicron neutralization capacity (adjusted OR (95% CI): 0.20 (0.07-0.53), 0.06 (0.01-0.24) and 0.04 (0.01-0.15) for previous infection, breakthrough infection and reinfection, respectively; *p*<0.001, Supplementary table 5). Males had higher odds to lose this ability (adjusted OR (95% CI): 2.64 (1.12-6.56); *p*=0.030, Supplementary table 5). Compared to the participants aged<40 years, those aged 40-64 years and ≥65 years had higher odds to lose Omicron neutralization capacity (adjusted OR (95% CI): 5.74 (1.87-22.19) and 7.89 (2.02-36.13), respectively; *p*=0.008, Supplementary table 5). Importantly, ethnic community was not associated with the loss of Omicron neutralization capacity between D3.1 and D3.6 (Supplementary table 5).

ADCC activation remained stable between D3.1 and D3.6 (median (95% CI) difference (+0.20 AU (-0.41; 0.61)). The multivariate linear regression model showed that the capacity to mediate ADCC in female participants who were never infected remained stable in time (mean (95% CI) progression -0.16 (-0.53; 0.21)) (Supplementary table 6). In participants with breakthrough infection between D3.1 and D3.6, the capacity to mediate ADCC significantly increased between D3.1 and D3.6 (mean (95% CI) progression of +0.51 AU (0.13; 0.88), *p*=0.0087). In male participants, the capacity to mediate ADCC significantly decreased between D3.1 and D3.6 (mean (95% CI) progression of -0.47 AU (-0.85; -0.08), *p*<0.018)). Importantly, the ethnic community was not associated with an evolution in the capacity to mediate ADCC (*p*=0.80 in univariate linear regression).

### Humoral response to SARS-CoV-2 at 6 months post-third dose is similar across ethnic communities

Among the 488 participants sampled at D3.6, 119 self-declared as Melanesians, 166 as Europeans, 68 as Polynesians and 135 belonged to other communities (Supplementary table 7). Europeans were older, had lower BMIs, exhibited signs of previous infection less frequently and were less able to neutralize Omicron. Past infection was significantly more frequent in Melanesians and “other communities” (*p*<0.001, Khi² test). Previous infection was significantly associated with higher anti-S IgG levels (crude effect +1.16, 95% CI (0.91; 1.41), *p*<0.001, Supplementary table 8). Melanesians and other communities showed higher anti-S IgG levels in univariate analysis. In multivariate analysis, only the infection status remained significantly associated with higher anti-S IgG levels.

Of these participants, 362 (74.2%) showed ability to neutralize Omicron. In the multivariate model, previous infection and higher anti-S IgG levels were independently associated with higher odds of neutralization at D3.6 (adjusted OR (95% CI): 4.04 (2.54-6.50), *p*<0.001 and 51.64 (11.00-922.94), *p*<0.001, respectively; Supplementary table 9). Conversely, males had lower odds to neutralize Omicron (adjusted OR (95% CI): 0.61 (0.38-0.97), *p*=0.024). As compared to participants aged <45 years, those aged 40-64 years and ≥65 years also had lower odds to neutralize Omicron (adjusted OR (95% CI): 0.48 (0.24-0.90) and 0.29 (0.14-0.57), *p*=0.0024). Capacity to neutralize Omicron at D3.6 was independent of the ethnic community.

## Discussion

This study found that Europeans, Melanesians, Polynesians and other communities had equivalent anti-S IgG titers and capacity to neutralize Omicron and mediate ADCC following Pfizer BNT162b2 vaccination and SARS-CoV-2 infection. They showed a similar maintenance of their hybrid humoral response 6 months after immunization. Melanesians’ and other communities’ higher anti-S IgG levels at D3.6 were associated with more frequent infections, probably resulting from different lifestyles across communities in NC, which may favor transmission ^2^.

Population genetic studies have revealed that ancient admixture between humans and Denisovans left traces in present-day Pacific Islanders’ genomes ^15^, introducing genetic variants almost exclusively related to immune response regulation ^16^. How genetic traits from archaic humans contribute to phenotypic differences between contemporary individuals, especially relating to their immune response, is poorly known. We showed here that Pacific Islanders’ genetic uniqueness did not translate in differential hybrid humoral response to SARS-CoV-2. Although some genetic variants have been associated with antigen-specific IgG levels ^17^, neither their presence in Pacific populations nor their impact on anti-SARS-CoV-2 IgG levels have been explored. Our results are consistent with robust antibody response and neutralization after two doses of BNT162b2 vaccine in a cohort comprising 80 Pacific people ^11^. Such efficient hybrid humoral response of Pacific Islanders highlights the importance of pursuing vaccination efforts in this population at risk for severe COVID-19.

We found that hybrid immunity resulting from BNT162b2 vaccination and SARS-CoV-2 infection was highly prevalent in our cohort. Due to COVID-19 epidemiology in NC (Supplementary figure 1), infected participants sampled at D2.1 or later timepoints were likely infected by the Delta variant, or either by the Delta or Omicron variants, respectively. In our study, hybrid immunity translated in a robust anti-S IgG secretion with efficient Omicron neutralization capacity compared to non-infected vaccinees, in Pacific Islanders as well. A third dose of BNT162b2 vaccine resulted in Omicron neutralization in almost all participants, independently from a previous infection. Consistently, hybrid immunity was shown to boost antibody levels and poly-functionality regardless of the sequence of natural infection and vaccination ^18^, resulting in a significant and lasting protection against Omicron infection ^19^. Recall of memory B cells generated after the original infection contributes to this increase in variant-neutralizing antibodies after vaccination ^20^. Enhancement of variant-neutralizing capacity in infected vaccinees supports the relevance to vaccinate Pacific Islanders, who exhibit higher COVID-19 seroprevalences (*Huet et al., in prep*).

We observed that self-declared males had lower anti-S IgG titers at one month after immunization, which did not translate in differential neutralization capacity. Sex did not affect antibody decline between D3.1 and D3.6. However, the loss of Omicron neutralization capacity between D3.1 and D3.6 was more likely in males, resulting in lower odds of Omicron neutralization at D3.6. Furthermore, the capacity to mediate ADCC was enhanced at one-month post-immunization but decreased more rapidly in males. Consistently, males were shown to secrete lower antibody levels after two doses of BNT162b2^21^. However, anti-S IgG levels were shown to decline faster in males after infection ^22^. Higher antibody titers in females were observed for several vaccines ^23,24^, suggesting a differential effective vaccine dose according to sex. Overall, while several studies demonstrated an impact of sex on the humoral response^22,25^, the underlying mechanisms remain poorly characterized.

In addition, we found that age did not affect antibody decline between D3.1 and D3.6, probably due to significantly lower anti-S IgG levels in the ≥40 years age groups at one-month after immunization. Consistently, older individuals exhibit lower antibody levels after two doses of BNT162b2, with similar decline across ages ^21^. Individuals aged ≥40 years had lower odds of Omicron neutralization at D3.6. Age might therefore impact humoral response polyfunctionality. SARS-CoV-2 vaccination’s outcome correlates with signs of immunosenescence ^26^, probably contributing to this loss of Omicron neutralization in older individuals. This highlights the importance to offer booster doses to the elderly to prevent the loss of Omicron neutralization capacity, in Pacific Islanders as well.

In this study, higher BMIs were associated with higher anti-S IgG levels at one-month post-immunization, in turn associated with higher odds of Omicron neutralization. However, impact of BMI on humoral immunity to natural infection and BNT162b2 vaccination is controversial ^27–29^. Nonetheless, this result emphasizes the benefits of vaccinating obese individuals, who are overrepresented among Pacific Islanders.

The current study suffers limitations: first, the high rate of SARS-CoV-2 infections prevented to assess ethnicity’s impact on the humoral response to vaccination alone. Nonetheless, infection status was taken into consideration in multivariate models. As SARS-CoV-2 immunity worldwide is predominantly hybrid, characterization of a population’s immunity, independently of its infection status, is highly relevant to adapt Public Health strategies. Second, the current study relied on self-declared ethnicity, without genetic determination of ancestry. Although mixed marriages are common, participants had the opportunity to identify as “Other communities” including metis. Self-declared ethnicity was thus an appropriate surrogate to genetically-determined ancestry. Finally, the sample size was not reached, hindering to ascertain response equivalence between communities. However, the lack of community effect, despite different prevalence of obesity, strongly suggests that SARS-CoV-2 hybrid humoral response does not differ between communities.

Overall, by investigating the hybrid immune response of Pacific Islanders to SARS-CoV-2, our study sheds light on the health of a population neglected in human studies, opening the door to considering Pacific Islanders’ uniqueness in Health studies.

## Methods

### Ethics

The current study was conducted in compliance with the Declaration of Helsinki principles and recorded on clinicaltrials.gov (ID: NCT05135585). Samples came from participants who gave their written informed consent as part of this study. This study was approved by the “Comité de Protection des Personnes Ile-de-France III” (n° ID-RCB 2021-A01949-32, CPP n° 4003-I, November 9th 2021, Am8898-1-4003 January 17th, 2022, Am9503-2-4003 April, 4th 2022 and Am9508-3-4003 June 21^st^, 2022) and by the Consultative Ethics Committee of New Caledonia.

### Cohort and study design

Enrolment took place in immunization centres in the South Province of NC, where adults (≥18 years) who received the Pfizer BNT162b2 vaccine were approached. Initially, individuals who received their second dose were approached. When the 3^rd^ dose was rolled out, participants who received their 3^rd^ dose were approached. In a third phase, individuals having received a 3^rd^ dose within the last 6 months were also approached (Figure 1). Documented SARS-CoV-2 infection (serologic, PCR or antigenic testing) was an exclusion criterion. Pregnant or breast-feeding women, immunosuppressed individuals, individuals unable to answer the questionnaire and those under protective measures were excluded. Sample size calculation for this equivalence study was based on a power of 85% to detect changes of more than 5% in the fraction of participants developing anti-S IgG in response to Pfizer BNT162b2 vaccination, with an alpha risk of 5%, resulting in the requirement to enroll 141 participants in each community.

Clinico-epidemiological data were recorded at enrolment in RedCap (Vanderbilt University, USA). A home visit by a qualified nurse was arranged one month (+/-10 days) after the second or third dose and either 3 months after the second dose or 6 months after the third dose. Upon these visits, a 4-mL venous blood sample was collected. Not all participants enrolled in the study participated to the follow-up. Sera were separated from blood samples by centrifugation and stored at -20°C until subsequent analyses.

### Cells and reagents

HEK 293T-hACE2 cells were cultured at 37°C under 5% CO_2_ in complete culture medium composed of Dulbecco’s modified Eagle’s medium (DMEM, Gibco) complemented with 10% fetal calf serum (Gibco), 1% penicillin/streptomycin (Gibco) and 2mM L-Glutamine (Gibco). Cells were detached and resuspended using 0.05% Trypsin-EDTA (Gibco).

Recombinant SARS-CoV-2 Wuhan Spike or Nucleocapsid proteins were kindly provided by S. Petres ^30^. Secondary lama antibodies VHH anti-human IgG Fc coupled to NanoKAZ were kindly provided by Thierry Rose. Z108 NanoKAZ substrate was kindly provided by Yves Janin.

### Pseudotyped viruses

Pseudotyped lentiviral vectors enveloped with the Spike protein from Wuhan, Delta, Omicron BA.1 or Omicron BA.4/5 SARS-CoV-2 variants were produced as previously described ^29^. Briefly, HEK 293T cells were co-transfected with a packaging plasmid Gag-Pol (NR-52517, BEI Resources), Helper Plasmids Tat1b (NR-52518, BEI Resources) and Rev1b (NR-52519, BEI Resources), a transfer plasmid carrying the gene encoding the Spike Glycoprotein of Wuhan-Hu-1 (GenBank: NC_045512, plasmid NR-52514, BEI Resources), Delta, Omicron BA.1 or Omicron BA.4/5 SARS-CoV-2 variants and a Lentiviral Backbone carrying a luciferase reporter gene Luc2 and a ZsGreen gene (pHAGE-CMV-Luc2-IRES-ZsGreen-W, NR-52516, BEI Resources). At 48h post-transfection, supernatants were collected, clarified by centrifugation and frozen at -80°C.

### LuLISA experiments

Sera were decomplemented at 56°C for 35 min. White 384-well tissue-culture plates (Greiner Bio-one) were coated with recombinant SARS-CoV-2 Wuhan Spike or Nucleocapsid proteins at 1 µg/mL in Phosphate Buffer Saline (PBS, Gibco) through 4h incubation at room temperature. Plates were washed with PBS containing 0.1% Tween (Tween®20, Sigma-Aldrich) using a plate washer Zoom microplate Washer (Titertek Berthold). Sera were diluted 1/200 in PBS containing 1% skimmed milk and 0.1% Tween using a Tecan Fluent 780 and incubated in duplicate for 2h in Spike- or Nucleoprotein-coated plates. Plates were washed with PBS containing 0.1% Tween. Secondary VHH lama antibodies anti-human IgG Fc coupled to NanoKAZ were diluted at 20 ng/mL in PBS containing 1% skimmed milk and 0.1% Tween and added to the plates. After 1h30 incubation, Z108 NanoKAZ substrate was added to the plates at 6 µg/mL. Bioluminescence was measured using Mithras² LB943 Multimode reader luminometer (Berthold technologies). Presence of anti-Spike IgG above the 0.25 AU was indicative of an immunization against SARS-CoV-2. Past COVID-19 infection was determined by the presence of anti-Nucleocapsid antibodies above 0.3 AU.

### Pseudovirus neutralization assay

Decomplemented sera (56°C for 35 min) were diluted 1/40 and coincubated with pseudotyped lentiviral vectors at room temperature for 50 min in complete culture medium prepared using DMEM without phenol red (Gibco). The mixture was then plated in white 96-well tissue-culture plates (Greiner Bio-One). A suspension of 20,000 HEK 293T-hACE2 cells was added in each well. Single-cell suspensions were prepared in Dulbecco’s Phosphate Buffer Saline (DPBS) containing 0.1% EDTA (Promega) to preserve hACE2 protein integrity. Serum-lentivirus mixes and addition of the cells were all performed using a Tecan Fluent 780. After 72h, Bright-Glo Luciferase substrate (Promega) was added to the cells (1:1). Bioluminescence was measured using Centro XS3 LB 960 or Mithras² LB943 Multimode reader luminometers (Berthold technologies).

A set of negative sera was used to determine the threshold as the average luminescence level of these negative sera minus 3 standard deviations.

The percentage of neutralization was expressed as (1 - (measured value/threshold)) x 100. The ability to neutralize Omicron variant was defined in pseudovirus neutralization assays as ≥90% neutralization of Omicron BA.1 and/or Omicron BA.4/5 pseudoviruses by a 1/40 dilution of the serum (ie neutralization titer >40).

### Antibody-Dependent Cellular Cytotoxicity (ADCC)

CD16 activation was quantified as a surrogate assay to measure sera’s capacity to mediate ADCC. ADCC was quantified using the ADCC Reporter Bioassay (Promega) and 293T cells stably expressing the Wuhan spike. Briefly, spike-expressing cells (3×10^4^ per well) were co-cultured with Jurkat-CD16-NFAT-rLuc cells (3×10^4^ per well) in presence or absence of sera diluted 1/30. Luciferase was measured after 18h of incubation using an EnSpire plate reader (PerkinElmer). ADCC was measured as the fold induction of Luciferase activity compared to the ‘‘no serum’’ condition. For each serum, the control condition (cells transfected with an empty plasmid) was subtracted to account for inter-individual variations of the background. We previously reported correlations between the ADCC Reporter Bioassay titers and an ADCC assay based on primary NK cells and cells infected with an authentic virus^13^. ADCC analyses were conducted on all samples collected at one-month post-immunization but two and on all corresponding follow-up samples collected 6 months after the 3^rd^ dose.

### Statistical analyses

Quantitative variables were characterized by the median, inter-quartile range (IQR) and range. Age was categorized into three groups: 18-39, 40-64 and ≥65 years. Participants with a Body Mass Index (BMI) strictly below 18.5 kg/m², comprised between 18.5 and 24.9 kg/m², comprised between 25 and 29.9 kg/m² or over 30 kg/m² were considered as underweight, normal weight, overweighted or obese, respectively.

Proportions were compared using Chi-2 test. Continuous variables were compared using Kruskal-Wallis test. The effect of immunization dose, infection status, self-declared gender, age, BMI category, presence of comorbidities and self-declared ethnicity on the levels of anti-Spike antibodies and CD16 activation was investigated using linear regression models. Their effect on Omicron neutralization capacity was investigated using logistic regression models. In all analyses, factors associated with the outcome with a *p*-value <0.20 in univariate analyses were considered in the multivariate model. Then, a backward stepwise procedure was used to identify factors that remained independently associated with the outcome. A *p*-value < 0.05 was considered significant. All statistical analyses were performed using R software (version R 4.2.1 GUI 1.79 High Sierra build (8095)).

## Data Availability

Pseudonymized data underlying the results presented in this study are available from the corresponding author, Catherine Inizan. Requests for reagents and resources should be directed to and will be fulfilled by the corresponding author, Catherine Inizan.

## Notes

### Author contributions

CI, MDR and VAD conceived the study. CI and MDR led the applications for funding with input from AC, GM, MJ and VAD. Consent and questionnaire tools were designed by CI and MDR. Inclusions were implemented by CI, AC, AFG, GM, VAD and MDR. Samples were collected by home-visiting nurses and handled by AB, ACG and AFG. CI, CS, AB, TB, VE, SM, OS and SW performed the experimentations. CI, AC and TD implemented the statistical analyses under the supervision from YM. CI wrote the scientific manuscript with input from YM, MDR and all authors. All authors have had the opportunity to access the underlying data used in this study. All authors reviewed the manuscript and approved the final version prior to submission.

## Acknowledgements

We would like to thank all the participants to the COVCAL study. This study has been labeled as a National Research Priority by the National Orientation Committee for Therapeutic Trials and other researches on COVID-19 (CAPNET). The investigators would like to acknowledge ANRS | Emerging infectious diseases for its scientific support and funding, the French Ministry of Health and Prevention and the French Ministry of Higher Education, Research and Innovation for their funding and support. We warmly thank the Clinical Research Department of the Centre for Translational Research at Institut Pasteur in Paris for their support in ethic procedures. The authors would also like to thank Priscillia Piersanti, Aurore Martini, Sabine Crescentini, Dr Christophe Assié and Dr Johanna Read along with home-visiting nurses and personnel from Medical Centers for their support in the investigation, Stéphane Petres and the Plateforme Technologique de Production et Purification de Protéines Recombinantes at Institut Pasteur for the production of the N and S proteins used in the LuLISA assays, Faustine Amara, Margot Penru, Yannis Rahou and Lou-Léna Vrignaud for technical support and Dr Etienne Simon-Lorière and Pr John Aaskov for insightful discussions.

## Data sharing

Further information and requests for reagents and resources should be directed to and will be fulfilled by the corresponding author, Catherine Inizan.

## Supplementary information titles and legends

**Supplementary Figure 1. Distribution of samples according to COVID-19 epidemiology in New Caledonia.** Absolute number of cases are shown in black on the left y axis, with Delta, Omicron BA.1 and Omicron BA.4/5 epidemic peaks. Absolute number of samples collected one month after the second dose (orange), three months after the second dose (green), one month after the third dose (blue) or 6 months after the third dose (violet) are shown on the right y axis. Inclusion periods are shown below the graph.

**Supplementary Figure 2. ROC curve for the anti-S IgG threshold best discriminating samples exhibiting Omicron neutralization capacity.**

**Supplementary Table 1.** Participants’ description at baseline (1 month after the 2^nd^ or 3^rd^ dose)

**Supplementary Table 2.** Comparison of immune characteristics after the 2^nd^ or 3^rd^ dose

**Supplementary Table 3.** Description of the participants followed up at month 1 and month 6 post-3^rd^ dose of vaccination (N=214)

**Supplementary Table 4.** Factors associated with the progression of anti-S IgG levels between one and six months after immunization (linear regression)

**Supplementary Table 5.** Factors associated with the loss of the ability to neutralize Omicron BA.1 and/or BA.4-5 between one and six months after immunization (logistic regression)

**Supplementary Table 6.** Factors associated with the variation in the level of ADCC (CD16 activation) between one and six months after immunization (linear regression)

**Supplementary Table 7.** Description of participants sampled at 6 months after the 3^rd^ dose of immunization

**Supplementary Table 8.** Factors associated with the levels of anti-S IgG six months after the 3^rd^ dose (linear regression)

**Supplementary Table 9.** Factors associated with the ability to neutralize Omicron BA.1 and/or BA.4-5 six months after the 3^rd^ dose (logistic regression)

